# Real-world Evidence of Diagnostic Testing and Treatment Patterns in U.S. Breast Cancer Patients with Implications for Treatment Biomarkers from RNA-sequencing Data

**DOI:** 10.1101/2020.08.07.20168401

**Authors:** Louis E. Fernandes, Caroline G. Epstein, Alexandria M. Bobe, Joshua S.K. Bell, Martin C. Stumpe, Michael E. Salazar, Ameen A. Salahudeen, Ruth A. Pe Benito, Calvin McCarter, Benjamin D. Leibowitz, Matthew Kase, Catherine Igartua, Robert Huether, Ashraf Hafez, Nike Beaubier, Michael D. Axelson, Mark D. Pegram, Sarah L. Sammons, Joyce A. O’Shaughnessy, Gary A. Palmer

## Abstract

**INTRODUCTION:** We performed a retrospective analysis of longitudinal real-world data (RWD) from breast cancer patients to replicate results from clinical studies and demonstrate the feasibility of generating real-world evidence. We also assessed the value of transcriptome profiling as a complementary tool for determining molecular subtypes.

**PATIENTS AND METHODS:** De-identified, longitudinal data were analyzed after abstraction from U.S. breast cancer patient records structured and stored in the Tempus database. Demographics, clinical characteristics, molecular subtype, treatment history, and survival outcomes were assessed according to strict qualitative criteria. RNA sequencing and clinical data were used to predict molecular subtypes and signaling pathway enrichment.

**RESULTS:** The clinical abstraction cohort (n=4,000) mirrored U.S. breast cancer demographics and clinical characteristics indicating feasibility for RWE generation. Among HER2+ patients, 74.2% received anti-HER2 therapy, with ~70% starting within 3 months of a positive test result. Most non-treated patients were early stage. In this RWD set, 31.7% of patients with HER2+ IHC had discordant FISH results recorded. Among patients with multiple HER2 IHC results at diagnosis, 18.6% exhibited intra-test discordance. Through development of a whole-transcriptome model to predict IHC receptor status in the molecular sequenced cohort (n=400), molecular subtypes were resolved for all patients (n=36) with equivocal HER2 statuses from abstracted test results. Receptor-related signaling pathways were differentially enriched between clinical molecular subtypes.

**CONCLUSION:** RWD in the Tempus database mirrors the overall U.S. breast cancer population. These results suggest real-time, RWD analyses are feasible in a large, highly heterogeneous database. Furthermore, molecular data may aid deficiencies and discrepancies observed from breast cancer RWD.

## Introduction

A growing number of studies have explored real-world data (RWD) and subsequent real-world evidence (RWE) to accelerate treatments for cancer patients. RWD relates to patient information procured during routine care, while RWE is the clinical evidence derived from RWD.^1-2^ The feasibility of this approach has increased alongside technological advances and regulatory support to continuously capture and integrate healthcare data sources.^3-7^ Several studies demonstrate the ability for RWE to guide clinical development strategies, expand product labels, and address knowledge gaps by examining clinical aspects not captured in clinical trials.^8-21^

An essential step towards strengthening RWE validity is demonstrating consistency between population statistics derived from observational RWD and those from controlled, experimental data. Despite the overwhelming support for RWE utility in oncology, technical barriers must be addressed for RWD/RWE to reach its full clinical potential. Incorporating administrative data, ancillary data, and unstructured clinical text from a variety of institutions to generate RWE is a complex task. For example, no standardization exists for abstracting and structuring highly heterogeneous data sources, and many natural language processing algorithms cannot account for these incongruencies.^2,3^ Consequently, clinical endpoints may not be accurately captured^22^ and even when data is properly abstracted and prepared for analysis, extraneous variables in raw RWD can introduce confounding biases.^7,23^ Similarly, the integration of omics data with RWD requires a controlled approach for large-scale data analytics.^24^

RWE and integrated omics data have the power to impact patient care.^3,25-27^ Various studies show the additive value of molecular tumor profiling with RWD for clinically relevant breast cancer insights,^8,28^ but further advancements in the field require the integration of genetic and clinical data from a variety of institutions, along with omics-focused capabilities and data analytics. One potential avenue to augment the value of breast cancer RWD is transcriptomics, as RNA-based gene expression analyses have shown prognostic, predictive, and treatment-directing value beyond DNA-sequencing insights.^29-36^ Whole-transcriptome RNA sequencing (RNA-seq) can help classify cancer types and breast cancer biomarkers,^37-39^ overcoming inconclusive pathology assessments, insufficient tissue quantity, and inter-observer variability of immunohistochemical or in-situ hybridization assays.^39-41^

Here, we address some of the complexities of RWD structuring and analyses. We demonstrate the feasibility of retrospective RWD analysis and test whether results from clinical studies can be replicated using longitudinal RWD from a large, representative breast cancer cohort. Our analyses present key clinical information, such as patient demographics, clinical characteristics, molecular markers, treatment patterns, and overall survival (OS) outcomes; and uncover discrepancies in real-world HER2 testing records. We also provide evidence supporting the integration of RWD with transcriptomic profiling for clinically relevant insights through analyses of RWD and molecular data from breast cancer patients sequenced by Tempus Labs.

## Patients and Methods

### Cohort Selections

Two retrospective breast cancer cohorts were randomly selected from the Tempus clinicogenomic database after applying clinically relevant inclusion criteria. All data were de-identified in accordance with the Health Insurance Portability and Accountability Act (HIPAA). Dates used for analyses were relative to the breast cancer primary diagnosis (pdx) date, and year of pdx was randomly off-set. Pdx within the cohorts spanned from 1990-2018. The first group was a clinical abstraction (CA) cohort of 4,000 breast cancer patients selected as a representative sample of RWD structured in the Tempus oncology database. Records were required to have data for a pdx, pdx date, age, race, sex, stage, histological subtype, and estrogen receptor (ER), progesterone receptor (PR), and human epidermal growth factor receptor 2 (HER2) status. The recorded stage and histological subtype were required to fall within 30 days relative to the pdx date, while the receptor statuses may have been recorded within 30 or 50 days, depending on the testing modality (see methods: *Molecular Subtype Determination*). A second cohort was selected, the Tempus molecular sequenced (MLC) cohort, which included 400 primary breast cancer patients with pdx dates and whose tumor biopsy underwent RNA-seq and targeted DNA sequencing (DNA-seq) with the Tempus xT (n=344), xO (n=55), or xE (n=1) panels between 2017-2019. While only patients with reported variants were included in the cohort, less than 1% of all breast cancer cases in the Tempus database have no DNA variants reported.

The study protocol was submitted to the Advarra Institutional Review Board. The IRB determined the research was exempt from IRB oversight and approved a waiver of HIPAA authorization for this study.

### Abstracted Molecular Markers

Protein expression from immunohistochemistry (IHC) results for ER and PR, as well as IHC and fluorescence in-situ hybridization (FISH) results for HER2 were curated during clinical data abstraction. Receptor results included abstracted equivocal, positive, or negative statuses. Hormone receptor (HR) status was classified by combinations of ER and PR statuses. When available, normalized Ki67 results included indeterminant, low, equivocal, moderate, or high statuses. A chi-squared test assessed the significance of Ki67 test result distribution differences. Fisher’s exact tests were performed for post-hoc analyses, and *P*-values were adjusted for multiple hypothesis testing using Bonferroni correction.

### Molecular Subtype Determination

The molecular subtype of each CA patient was classified as HR+/HER2-, HR+/HER2+, HR-/HER2+, or triple-negative breast cancer (TNBC) based on their receptor statuses at diagnosis. HR statuses were determined from the most recent IHC results or physician notes recorded within 30 days of the pdx date. HR+ status included ER+/PR+, ER+/PR-, and ER-/PR+. HER2 status was determined from the most recent FISH results recorded within 50 days of the pdx date. In the absence of HER2 FISH data, the most recent IHC result or physician note within 30 days of the pdx date was utilized. References to results at “initial diagnosis” imply these 30- and 50-day time frames. Molecular subtypes in the MLC cohort were determined from IHC or FISH results associated with the patient pathology report.

### Clinical Data Abstraction

Clinical data were extracted from the Tempus real-world oncology database of longitudinal structured and unstructured data from geographically diverse oncology practices, including integrated delivery networks, academic institutions, and community practices. Many of the records included in this study were obtained in partnership through ASCO CancerLinQ. Structured data from electronic health record systems were integrated with unstructured data collected from patient records via technology-enabled chart abstraction and corresponding molecular data, if applicable. Data were harmonized and normalized to standard terminologies from MedDRA, NCBI, NCIt, NCIm, RxNorm, and SNOMED.

### Menopausal Status Determination

Menopausal status was determined using relevant abstracted text fields when available. A patient was considered premenopausal if a single, undated menopause-negative (perimenopausal, premenopausal, or menstruating) status was recorded on or prior to the pdx date and no menopause-positive (menopausal or postmenopausal) status was indicated before diagnosis. Patients were also considered premenopausal at pdx if a menopausal event was recorded after a year from the pdx date.

Likewise, patients with an undated menopause-positive status, and patients with a menopausal or postmenopausal status recorded on or prior to the pdx date, were considered postmenopausal. A patient was also considered postmenopausal if no menopausal information was available on or prior to the pdx date, but a menopausal or postmenopausal status was indicated within one year after.

Menopausal status circumstances beyond the scope of these criteria were denoted as “Unknown.”

### Overall Survival Analysis

Overall survival (OS) was calculated for all stage I-IV CA cohort patients with invasive breast cancer (n=3,952). Patients without known relative death dates were right censored at their most recent relative clinical interaction date. Survival curves were generated in R using the survival (v2.43-4) and survminer (v0.4.3) packages with *P*-values calculated by log-rank tests. Results depict the percentage of surviving patients per year, and are stratified based on stage and HER2, ER, and triple-negative status.

### Genomic Testing

MLC cohort reported variants were generated from targeted DNA-seq of formalin-fixed, paraffin-embedded (FFPE) slides of primary breast tumor biopsies and, when possible, matched saliva or blood samples. Whole-transcriptome RNA-seq was performed on samples from the same tissue block. Most samples were sequenced with the Tempus xT or xO targeted DNA-seq assays, which detect oncologic targets in solid tumors and hematological malignancies as previously described.^37,42^ Two patient samples were sequenced with an updated and refined version of the xT panel targeting clinically relevant exons in 596 genes, and their reported variants were merged for analyses. Additionally, one sample in the MLC cohort was sequenced with the Tempus xE assay, a whole-exome panel targeting 19,396 genes over a 39 megabase (Mb) genomic region.

### Genomic Test Variant Reporting

Because each Tempus assay targets different gene sets, MLC cohort variant analyses only included genes tested across all 400 samples.^42,43^ Variants were classified and reported according to previously established clinical guidelines.^37^ Reported variants were categorized as alterations, fusions, or copy number variation amplifications or deletions. Alterations include variants of unknown significance (VUS), biologically relevant or potentially actionable alterations, and both germline VUS and pathogenic variants.

### Tumor mutational burden (TMB)

TMB was calculated by dividing the number of non-synonymous mutations by the adjusted panel size of the xT, xO, or xE assay (2.4 Mb, 5.86 Mb, and 36 Mb, respectively). All non-silent somatic coding mutations, including missense, indel, and stop-loss variants with coverage greater than 100x and an allelic fraction greater than 5% were counted as non-synonymous mutations.

### RNA-based Prediction of Molecular Subtypes

Transcriptome models were used to predict receptor statuses for the MLC cohort, including patients lacking IHC or FISH data. Briefly, single-gene logistic models were trained on an independent set of Tempus RNA-sequenced breast cancer samples according to the normalized gene expression of *ESR1*, *PGR*, or *ERBB2* using the R glm package v2.0-16. Model performances were assessed separately for primary samples, metastatic samples, and a combined set using 10-fold cross-validation (**Supplemental Table 2**). Performance was evaluated on a testing set comprised of RNA-sequenced samples in the MLC cohort with abstracted IHC or FISH results in the Tempus database (ER n=308, PR n=306, HER2 n=261). These samples were withheld from the training set. Positivity thresholds for IHC prediction models were selected using Youden’s J statistic to optimize sensitivity and specificity.

### Gene Expression Collection, Processing, and Normalization

Gene expression was generated through RNA-seq of FFPE tumor samples using an exome capture-based protocol.^37^ Transcript-level quantification to GRCh37 was performed using Kallisto 0.44. Transcript counts were then corrected for GC content and length using quantile normalization and adjusted for sequencing depth via a size factor method. Normalized counts in protein coding transcripts covered by the exome panel were then summed to obtain gene-level counts. Subsequent expression analyses were performed on log_10_-transformed counts.

### RNA-seq Pathway Analyses

Gene sets were downloaded from the MSigDB website (http://software.broadinstitute.org/gsea/msigdb/index.jsp), and pathway enrichment scores were calculated from normalized gene expression using the ssGSEA function in Gene Set Variation Analysis (GSVA) R Bioconductor package v1.0.6.^37,44^ ER- and HER2-related pathways were identified as those containing the terms “*ESR1*” or “Estrogen” and “*ERBB2*” or “HER2,” respectively. Z-scores were calculated for each set of enrichment scores and the sign was reversed for any pathway containing “DN” (down) or “repressed.” For select analyses, the mean of the z-score across pathways was calculated to produce a patient pathway metascore. With the exception of the HER2 and ER signaling pathway metascore analyses, receptor status was derived from both abstracted and predicted protein expression. Significance was determined by a Wilcoxon test for any comparison between two groups, and a Kruskal-Wallis test for comparisons between three or more groups, with *P*<0.05 considered significant. A separate gene set analysis was conducted to test the difference in enrichment among the four molecular subtypes relative to the 50 Hallmark pathways, a highly curated list from the MSigDB database.^45^ To determine how patients clustered by pathway scores, we performed a second UMAP analysis with enrichment scores for each Hallmark pathway as features.

## Results

### Real-world evidence from a clinical abstraction breast cancer cohort

#### Patient demographics and clinical characteristics in the CA cohort

We first determined whether key demographic and clinical characteristics captured in RWD replicate clinical studies, and found the deidentified data were consistent with previous large-scale breast cancer cohort studies (**Table 1**).^46-49^ The cohort mostly comprised females (99.3%, n=3,970) with a median age at diagnosis of 61.0 years. Year of diagnosis among the cohort ranged from 1990 to 2018 (**Supplemental Fig. 1**). The self-reported race was 83.3% White (n=3,332), 13.1% Black or African American (n=523), and 3.6% Asian or Pacific Islander (n=145). In 2,042 females with menopausal data, 87.4% (n=1,784) were postmenopausal. Abstracted stage at initial diagnosis primarily consisted of stage I (49.6%, n=1,986) and II (33.3%, n=1,333), followed by III (10.5%, n=420), IV (5.5%, n=219), and 0 (1.1%, 42). Most tumors had a histological classification of invasive ductal carcinoma (77.4%, n=3,095), and 9.5% (n=378) had an invasive ductal component or were NOS. Several rare cancer types were also represented.

**Table 1.**
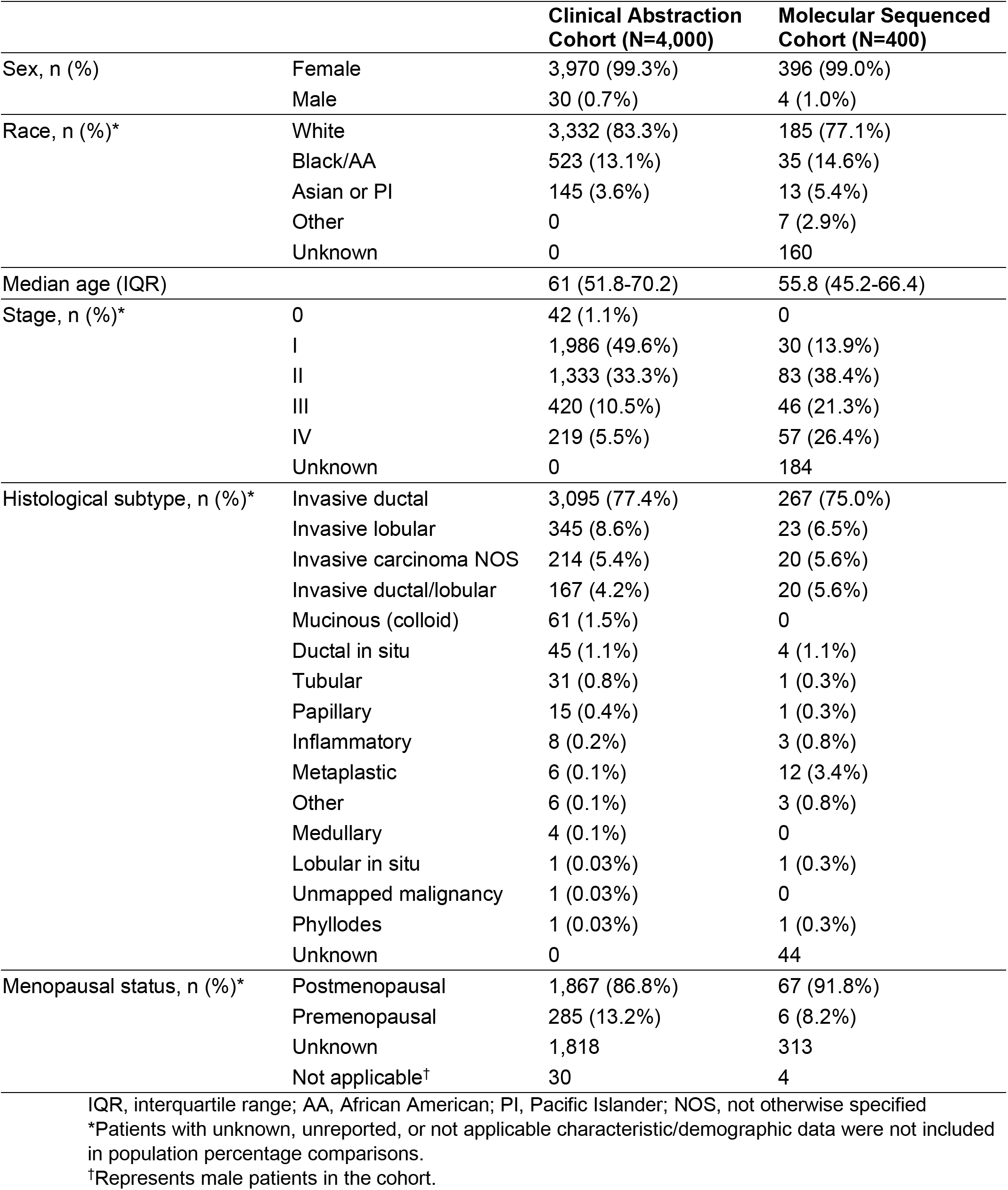
Demographics and clinical characteristics of the clinical abstraction and Tempus molecular sequenced cohort.

|  |  | **Clinical Abstraction Cohort (N=4,000)** | **Molecular Sequenced Cohort (N=400)** |
| --- | --- | --- | --- |
| Sex, n (%) | Female | 3,970 (99.3%) | 396 (99.0%) |
|  | Male | 30 (0.7%) | 4 (1.0%) |
| Race, n (%)* | White | 3,332 (83.3%) | 185 (77.1%) |
|  | Black/AA | 523 (13.1%) | 35 (14.6%) |
|  | Asian or PI | 145 (3.6%) | 13 (5.4%) |
|  | Other | 0 | 7 (2.9%) |
|  | Unknown | 0 | 160 |
| Median age (IQR) |  | 61 (51.8-70.2) | 55.8 (45.2-66.4) |
| Stage, n (%)* | 0 | 42 (1.1%) | 0 |
|  | I | 1,986 (49.6%) | 30 (13.9%) |
|  | II | 1,333 (33.3%) | 83 (38.4%) |
|  | III | 420 (10.5%) | 46 (21.3%) |
|  | IV | 219 (5.5%) | 57 (26.4%) |
|  | Unknown | 0 | 184 |
| Histological subtype, n (%)* | Invasive ductal | 3,095 (77.4%) | 267 (75.0%) |
|  | Invasive lobular | 345 (8.6%) | 23 (6.5%) |
|  | Invasive carcinoma NOS | 214 (5.4%) | 20 (5.6%) |
|  | Invasive ductal/lobular | 167 (4.2%) | 20 (5.6%) |
|  | Mucinous (colloid) | 61 (1.5%) | 0 |
|  | Ductal in situ | 45 (1.1%) | 4 (1.1%) |
|  | Tubular | 31 (0.8%) | 1 (0.3%) |
|  | Papillary | 15 (0.4%) | 1 (0.3%) |
|  | Inflammatory | 8 (0.2%) | 3 (0.8%) |
|  | Metaplastic | 6 (0.1%) | 12 (3.4%) |
|  | Other | 6 (0.1%) | 3 (0.8%) |
|  | Medullary | 4 (0.1%) | 0 |
|  | Lobular in situ | 1 (0.03%) | 1 (0.3%) |
|  | Unmapped malignancy | 1 (0.03%) | 0 |
|  | Phyllodes | 1 (0.03%) | 1 (0.3%) |
|  | Unknown | 0 | 44 |
| Menopausal status, n (%)* | Postmenopausal | 1,867 (86.8%) | 67 (91.8%) |
|  | Premenopausal | 285 (13.2%) | 6 (8.2%) |
|  | Unknown | 1,818 | 313 |
|  | Not applicable† | 30 | 4 |

IQR, interquartile range; AA, African American; PI, Pacific Islander; NOS, not otherwise specified
\*Patients with unknown, unreported, or not applicable characteristic/demographic data were not included in population percentage comparisons.
†Represents male patients in the cohort.

#### Molecular subtype determination in the CA cohort

We assessed the extent to which RWD captures molecular marker information from clinical testing results. The distributions of all abstracted receptor testing results at initial diagnosis are shown in **Fig. 1A**. Consistent with previous U.S. breast cancer statistics,^50^ the most prevalent molecular subtype was HR+/HER2- (71.5%, n=2,859), followed by TNBC (12.3%, n=491) (**Fig. 1B**). Among HR+ patients with non-equivocal statuses, most were ER+/PR+ (71.0%, 2,839 of 3,996) followed by ER+/PR- (10.4%, n=417) and ER-/PR+ (1.4%, n=57) (**Fig. 1C**). Lastly, abstracted Ki67 IHC test results were consistent with the Ki67 expression levels typically indicative of specific breast cancer subtypes (**Fig. 1D**).^51-52^ The distribution of Ki67 results differed significantly among molecular subtypes (chi-squared, *P*=1.75×10^-9^), particularly between HR+/HER2- versus HR+/HER2+ patients (*P*=0.015) and TNBC versus HR+/HER2- patients (*P*=6.38×10^-9^). The largest proportions of high Ki67 IHC test results were in TNBC (82.0%, n=50 of 61) and HR-/HER2+ patients (75.0%, n=15 of 20), while most low Ki67 results were in HR+/HER2- patients (44.0%, n=140 of 318).

**Figure 1.**
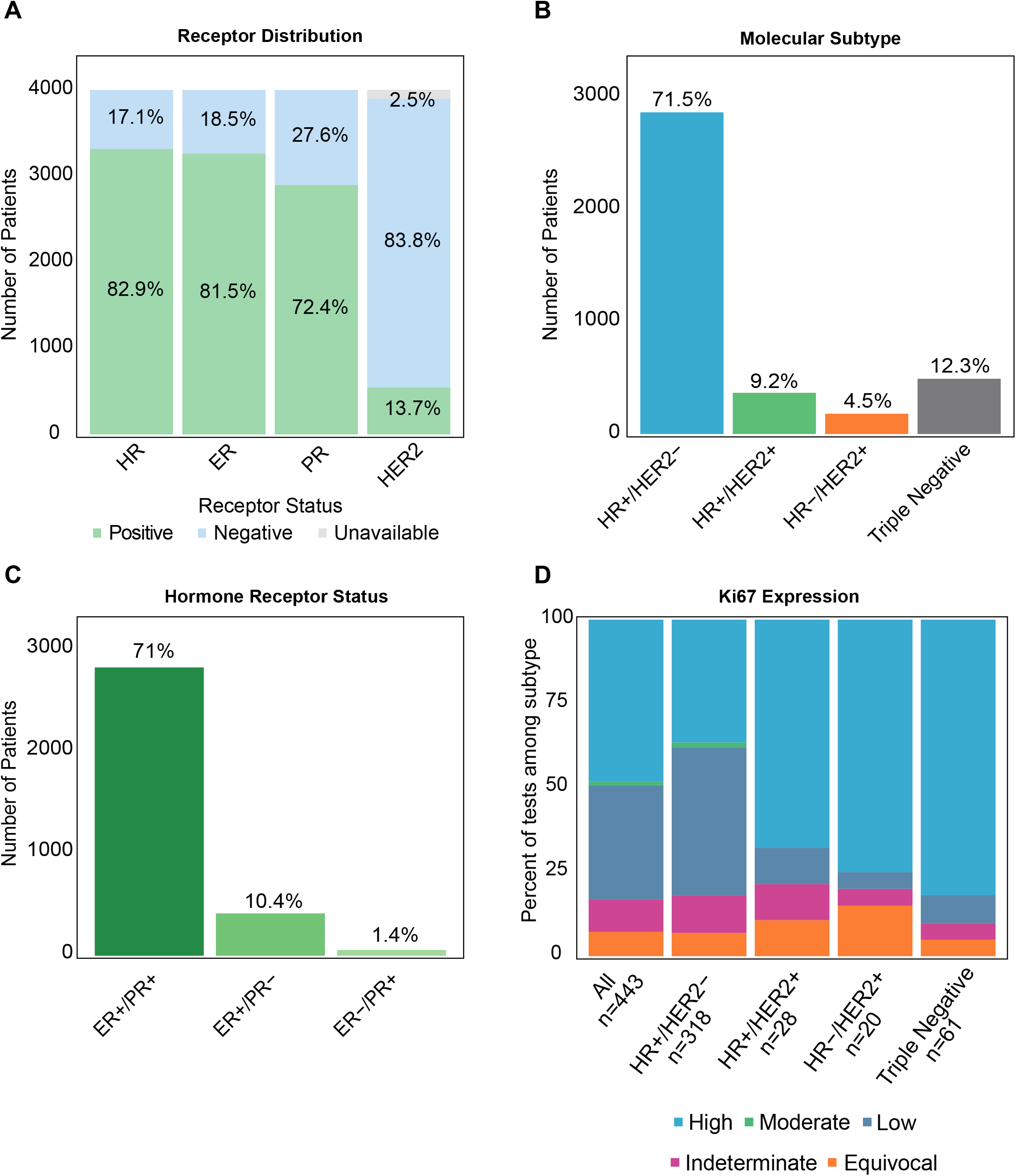
Breast cancer molecular biomarkers and subtypes in the clinical abstraction cohort. (A) The number of patients with positive, negative, or equivocal IHC or FISH test results for ER, PR, HR, and HER2 status at initial diagnosis. (B) The distributions of breast cancer molecular subtypes as determined by abstracted ER, PR, and HER2 test results at initial diagnosis, and (C) the distribution of ER and PR status combinations across the cohort. (D) The number of patients with high, moderate, low, indeterminate, or equivocal Ki67 IHC test results or status-indicating physician notes at initial diagnosis, separated by molecular subtype.

#### Anti-HER2 therapy analysis in the CA cohort

We next examined anti-HER2 therapy treatment patterns from longitudinal RWD. Curated anti-HER2 therapies included trastuzumab, ado-trastuzumab emtansine, neratinib, lapatinib and pertuzumab. Among CA cohort patients, 13.7% (n=546) were HER2+ at initial diagnosis, of whom 74.2% (n=405) received anti-HER2 therapy at some point in their clinical care. Approximately 70.0% of patients who received anti-HER2 therapy did so within 3 months of a positive test result and the majority (73.5%) had early-stage cancer (**Fig. 2A**). These results are consistent with previous breast cancer cohort studies.^16,53^ Moreover, a small portion of HER2- patients exhibited evidence of receiving an anti-HER2 therapy (1.1%, 36 of 3,352 HER2- patients) (**Fig. 2B**). Of those patients, 33.3% (n=12) had evidence of a discordant result at initial diagnosis, 44.4% (n=16) had only HER2- results, and 22.2% (n=8) had a HER2-equivocal or positive result recorded beyond initial diagnosis. A small portion of patients (n=37) were not assigned a HER2 treatment time frame due to date quality issues.

**Figure 2.**
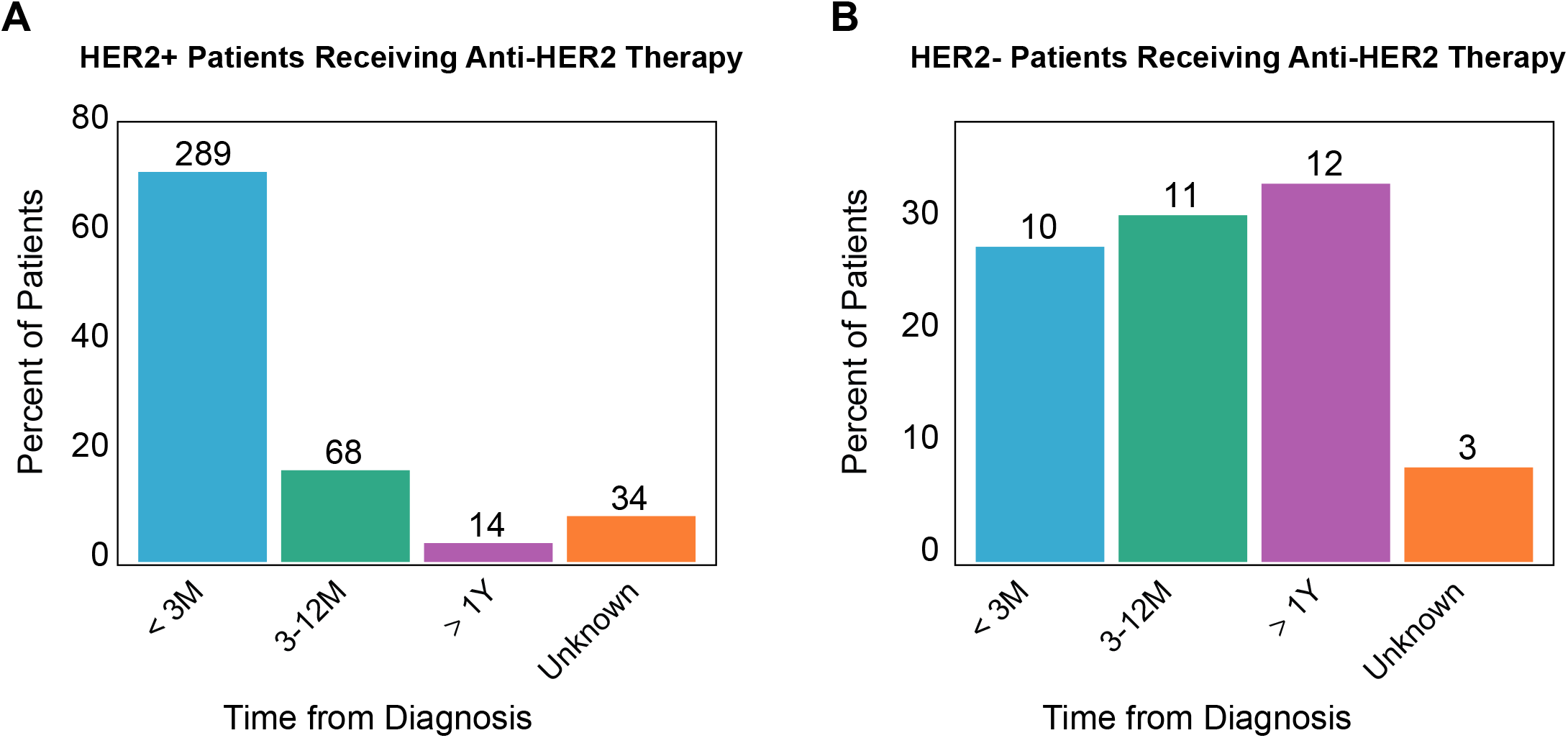
Anti-HER2 treatment by HER2 status in the clinical abstraction cohort. Anti-HER2 treatment initiation patterns among (A) HER2+ and (B) HER2- patients who received anti-HER2 therapy at some point in their clinical care. M, month; Y, year.

#### HER2 test result analyses in the CA cohort

To evaluate inter- and intra-test concordance, we compared HER2 IHC and FISH results among patients with both tests conducted near initial diagnosis (17.7%, n=709). Among patients with HER2+ IHC results and subsequent FISH testing, 62.2% (n=51 of 82) were inter-test concordant (**Supplemental Table 1**), however, 31.7% with HER2+ IHC were HER2- by FISH (n=26 of 82). This discordance is larger than a previously reported meta-analysis of IHC and FISH HER2 testing worldwide.^54^ Four of those 26 patients had received an anti-HER2 therapy in their clinical timeline. Among patients with HER2-IHC results, 3.9% (n=7 of 182) were HER2+ by FISH, similar to historical reports.^54^ The majority of these patients (n=6 of 7) received anti-HER2 therapy. HER2-equivocal IHC results (HER2 IHC 2+) were observed in 62.8% (n=445 of 709) of the cohort. Among these patients with equivocal results, 7.8% (n=35 of 445) were later confirmed equivocal by FISH testing. However, 80.7% (n=359 of 445) had subsequent HER2- and 11.5% (n=51 of 445) HER2+ FISH results.

Additionally, intra-test discordance was analyzed in patients with multiple HER2 results at initial diagnosis. Among patients with multiple HER2 IHC results at diagnosis (7.1%, n=253 of 3,561 with HER2 IHC), 18.6% (n=47) exhibited intra-test discordance. Of patients with multiple HER2 FISH results (4.5%, n=52 of 1,157), 21.2% (n=11) exhibited intra-test discordance.

#### Overall survival in the CA cohort

OS analyses from longitudinal RWD revealed overall 5-year and 10-year survival rates (92.2% and 85.7%, respectively) relatively consistent with average U.S. percentages (**Fig. 3A**). ^46^ Survival rates were expectedly high, varying by stage (*P*<0.0001) and receptor status. Stage IV patients exhibited worse OS than earlier-stage patients (**Fig. 3B**). The 5-year survival rate was 93.5% in stage I-IV HER2+ patients and 92.0% in HER2-patients (*P*=0.45), with rates of 74.3% and 57.1%, respectively, among stage IV patients (*P*=0.098) (**Fig. 3C, 3D**). The 5-year survival rate was 92.7% among stage I-IV ER+ patients and 89.8% in ER-patients (*P*=0.052), with rates of 63.7% and 55.5%, respectively, among stage IV patients (*P*=0.12) (**Fig. 3E, 3F**). TNBC patients had significantly worse OS compared to other subtypes, with a 36.3% 5-year survival rate in stage IV TNBC patients compared with 65.1% among stage IV non-TNBC patients (*P*=0.0024) (**Fig. 3G, 3H**).

**Figure 3.**
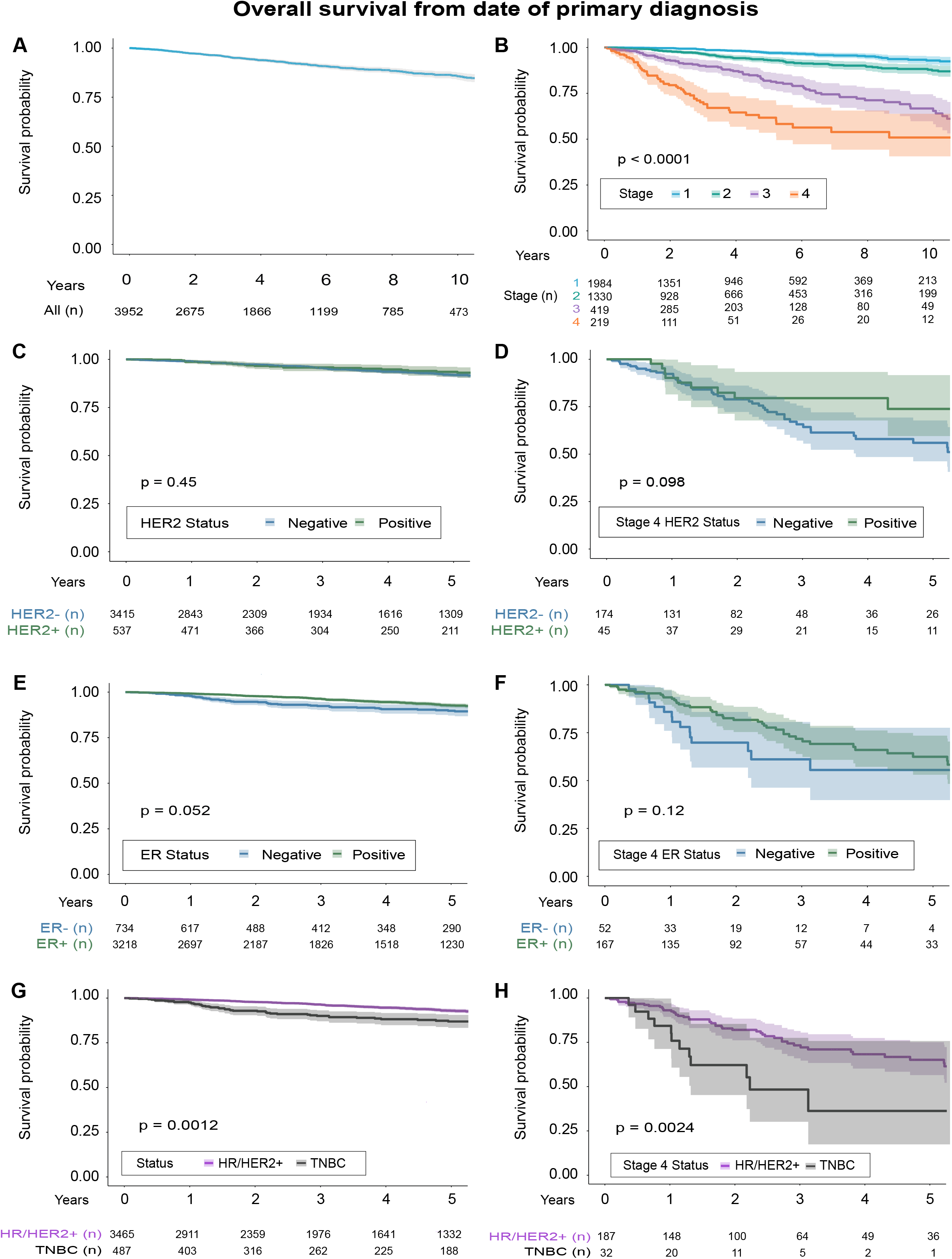
Overall survival from primary diagnosis dates in the clinical abstraction cohort. Ten-year survival probability in (A) all patients and (B) stage I-IV patients stratified by stage. Five-year survival probabilities stratified by HER2 status in (C) all patients and (D) stage IV patients, ER status in (E) all patients and (F) stage IV patients, and TNBC status in (G) all patients and (H) stage IV patients.

### Genomic testing insights from the Tempus molecular sequenced cohort

#### Patient demographics and clinical characteristics in the MLC cohort

Abstracted clinical characteristics and patient demographics from the 400 MLC cohort patients were assessed (**Table 1**), and found to be relatively consistent with the CA cohort and other large-scale breast cancer cohort studies.^46-49^ The cohort had a slightly younger median age at diagnosis of 55.8 years (45.2-66.4), and higher percentage of Black or African American (14.6%, n=35) and Asian or Pacific Islander patients (5.4%, n=13) than the CA cohort. Patients with known stage information were mostly stage II at diagnosis (38.4%, n=83), followed by stages IV (26.4%, n=57), III (21.3%, n=46), and I (13.9%, n=30), indicating an overall higher risk population compared with the CA cohort. A total of 75.0% (n=267) of tumors were invasive ductal carcinoma, with several rare cancer types also represented in the cohort.

#### DNA sequencing analysis of the MLC cohort

The top three genes with reported alterations were *TP53*, *PIK3CA*, and *GATA3*, which were found in 55.0% (n=220), 29.0% (n=116), and 13.8% (n=55) of the MLC cohort, respectively (**Supplemental Fig. 2A**). These findings are consistent with a previous analysis of The Cancer Genome Atlas breast cancer data.^55^ **Supplemental Fig. 2B** shows the distribution of variant types in the 20 most frequently reported genes. Assessment of patients with tumor/normal-matched DNA-seq (n=356) identified 18 patients (5.1%) with pathogenic germline variants in 12 NCCN-designated familial high-risk genes (**Supplemental Fig. 2C**). This sub-population may be underrepresented as exon-level duplications or deletions were not included. Among the 18 patients harboring a pathogenic germline variant in any of those 12 genes, most contained variants in *BRCA 1* or *2* (n=11), followed by *CHEK2* (n=6), *ATM* (n=3), and *PALB2* (n=2). Because TMB and MSI status are integrated biomarker measurements in the Tempus platform, we observed a wide range of TMB across the cohort with a median of 1.7 mutations/Mb (**Supplemental Fig. 2D**). Consistent with previous studies,^56^ the majority of patients (84.7%, n=339) were MSI stable, while only 0.3% (n=1) were MSI high and 0.5% (n=2) were MSI low.

#### RNA-based prediction of receptor status for molecular subtypes

We developed a whole-transcriptome model based on 19,147 genes to predict IHC receptor status and resolve molecular subtypes in the MLC cohort. Predicted RNA-based subtypes largely aligned with abstracted IHC-based subtypes (**Fig. 4A**). Similar to the literature,^57-58^ transcriptome signatures differed between molecular subtypes with TNBC clustering separately. Seventeen samples clustered with TNBC but were predicted or abstracted as another subtype, suggesting samples that cluster outside of their groups may benefit from further testing or analysis. *ESR1*, *PGR*, and *ERBB2* gene expression correlated with their respective abstracted and predicted receptor statuses (**Fig. 4B**).

**Figure 4.**
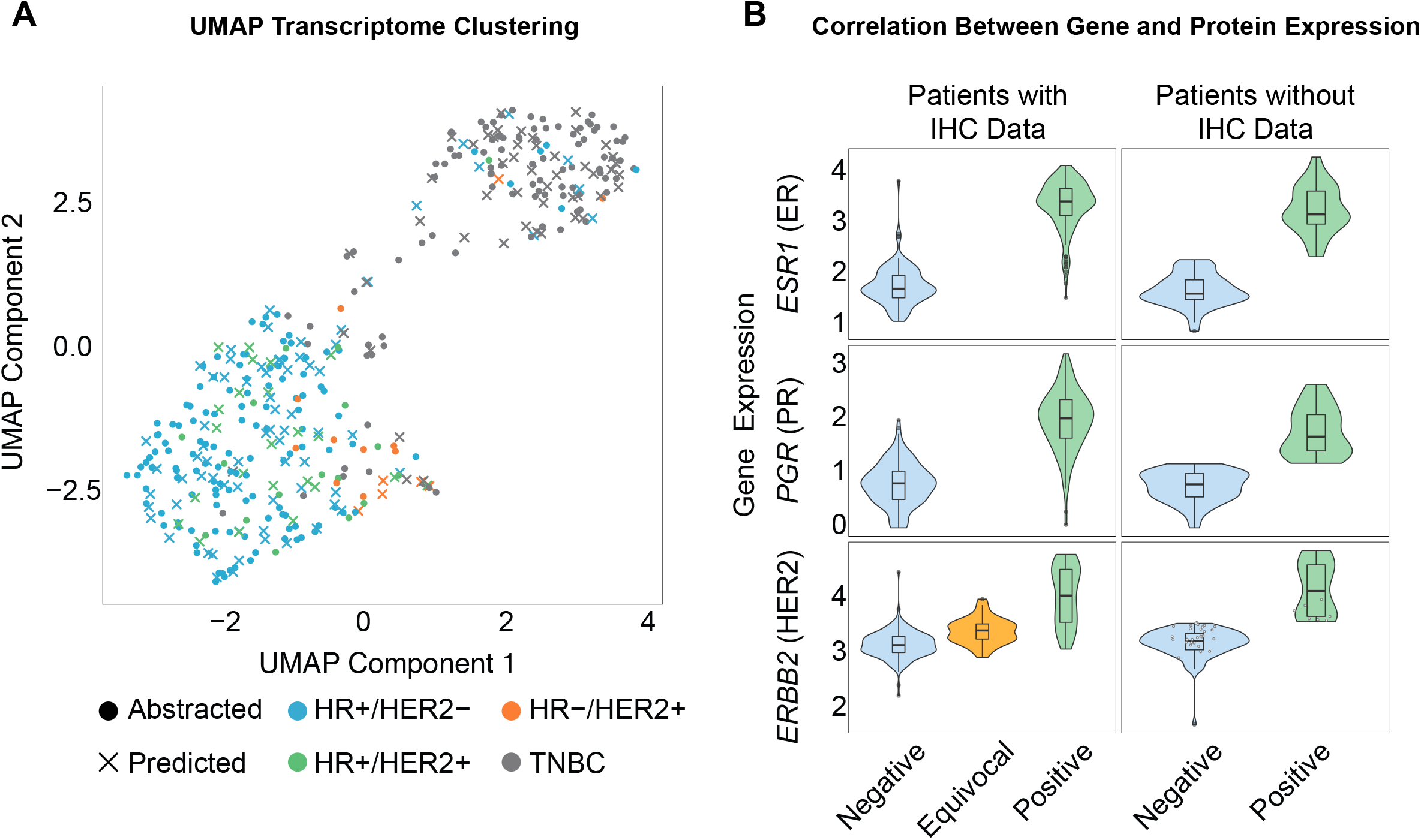
RNA-based receptor status prediction analysis of the Tempus molecular sequenced cohort. (A) UMAP transcriptome clustering of 19,147 genes in the cohort color-coded by molecular subtype. Circles correspond to samples with available IHC or FISH test results for all proteins and X symbols correspond to patients with predicted status for at least one protein. (B) Relationship between ER, PR, and HER2 receptor status and log_10_-transformed, normalized gene expression of *ESR1*, *PGR*, and *ERBB2*. Left panels represent samples with available receptor status from abstracted test results, while right panels represent transcriptome-based receptor status predictions. HER2 predictions for samples reported as equivocal are plotted as white dots.

RNA-based receptor status predictions were highly accurate for ER (95.5%, AUROC 98.1%) and HER2 (94.6%, AUROC 93.8%) relative to abstracted status, while PR status was predicted with slightly lower accuracy (87.9%, AUROC 95.2%) (**Fig. 5**). Prediction accuracy for all receptors was 92.7%. A detailed overview of the validation data and model performance are available in **Supplemental Table 2**. Patients with incompletely abstracted molecular subtypes (n=150) were classified by predicted receptor statuses from the transcriptomic model. Importantly, patients with equivocal HER2 statuses abstracted from IHC and/or FISH results (n=36) were predicted HER2+ (n=7) or HER2-(n=29) by the model.

**Figure 5.**
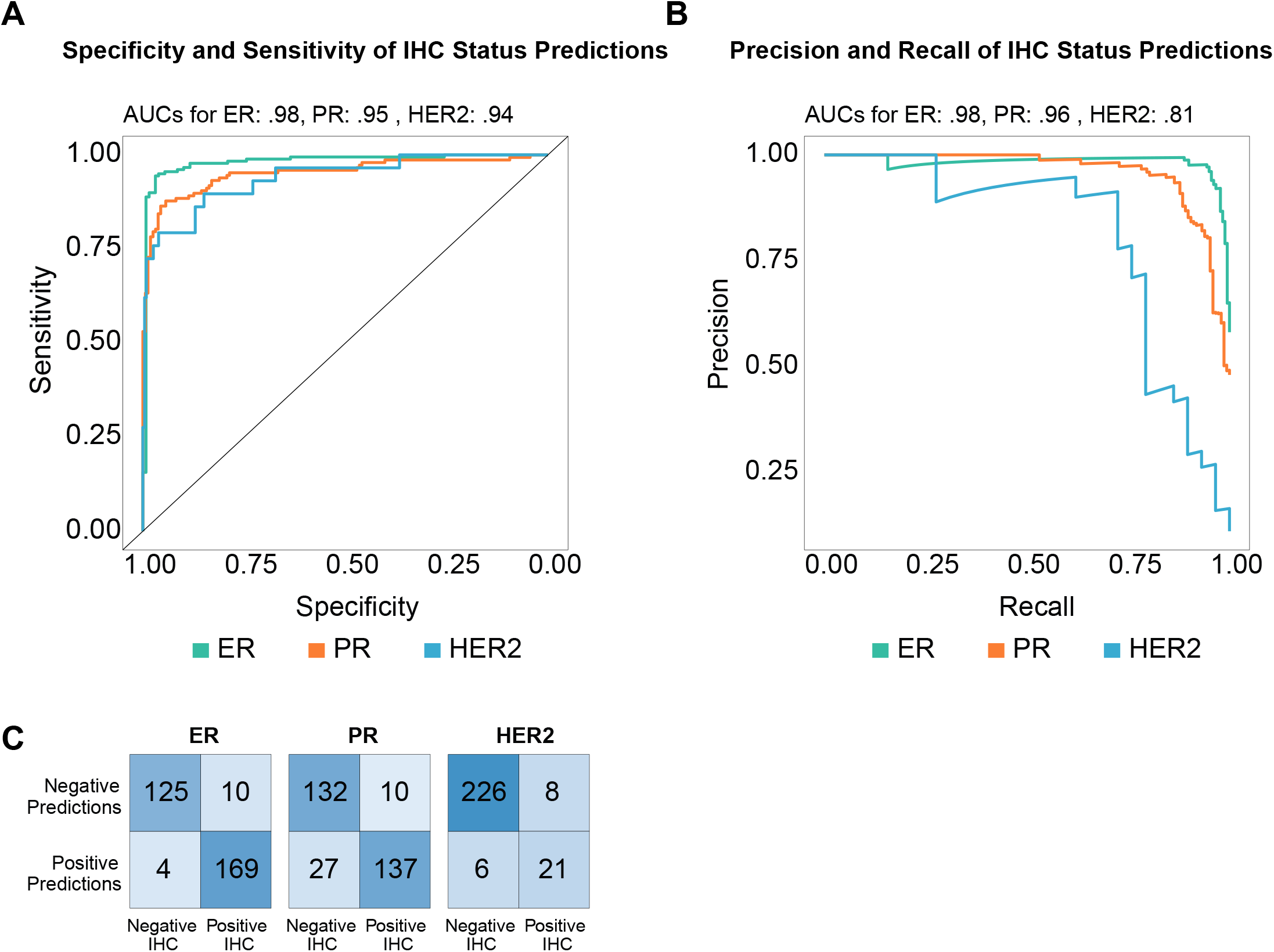
Single-gene logistic model performance for ER, PR, and HER2 status prediction in the Tempus molecular sequenced cohort. The (A) specificity and sensitivity, and (B) precision and recall of transcriptome-based receptor status predictions were evaluated on a testing set comprised of cohort RNA-sequenced samples with abstracted receptor status results in the Tempus database. (C) Confusion matrices depicting transcriptome-based ER, PR, and HER2 status prediction performance.

#### RNA-based HER2 and ER pathway analyses

To further evaluate the potential for RNA-seq to enhance breast cancer clinical data, a gene set enrichment analysis was conducted using the MSigDB database. First, we assessed whether measuring the activity of signaling pathways may resolve ambiguous or equivocal IHC and FISH test results. Multiple gene sets that putatively measure such pathway activity were identified by searching for “*ERBB2*,” “HER2,” “*ESR1*,” or “Estrogen” in the MSigDB database (**Supplemental Fig. 3A and 3B**). Results of the pathway analyses were expressed as metascores to avoid the bias introduced when selecting a single pathway. HER2 IHC-positive and FISH-positive samples were enriched for HER2 activity metascores as expected, but the HER2 signaling results contained substantial variability in pathway activity (**Fig. 6A**). Notably, the GO_ERBB2_SIGNALING_PATHWAY, which directly measures HER2 activity,^59^ exhibited a robust correlation with HER2 expression (r=0.453) (**Supplemental Fig. 3A)** and significantly different enrichments between HER2 statuses (*P*=0.00031) **(Supplemental Fig. 5**). While ER enrichment scores were more distinct between IHC-positive and IHC-negative patients, consistent with the relatively higher reliability of ER IHC compared with HER2 tests,^60-62^ variability was also observed in the ER signaling results (**Fig. 6B, Supplemental Fig. 6**).

**Figure 6:**
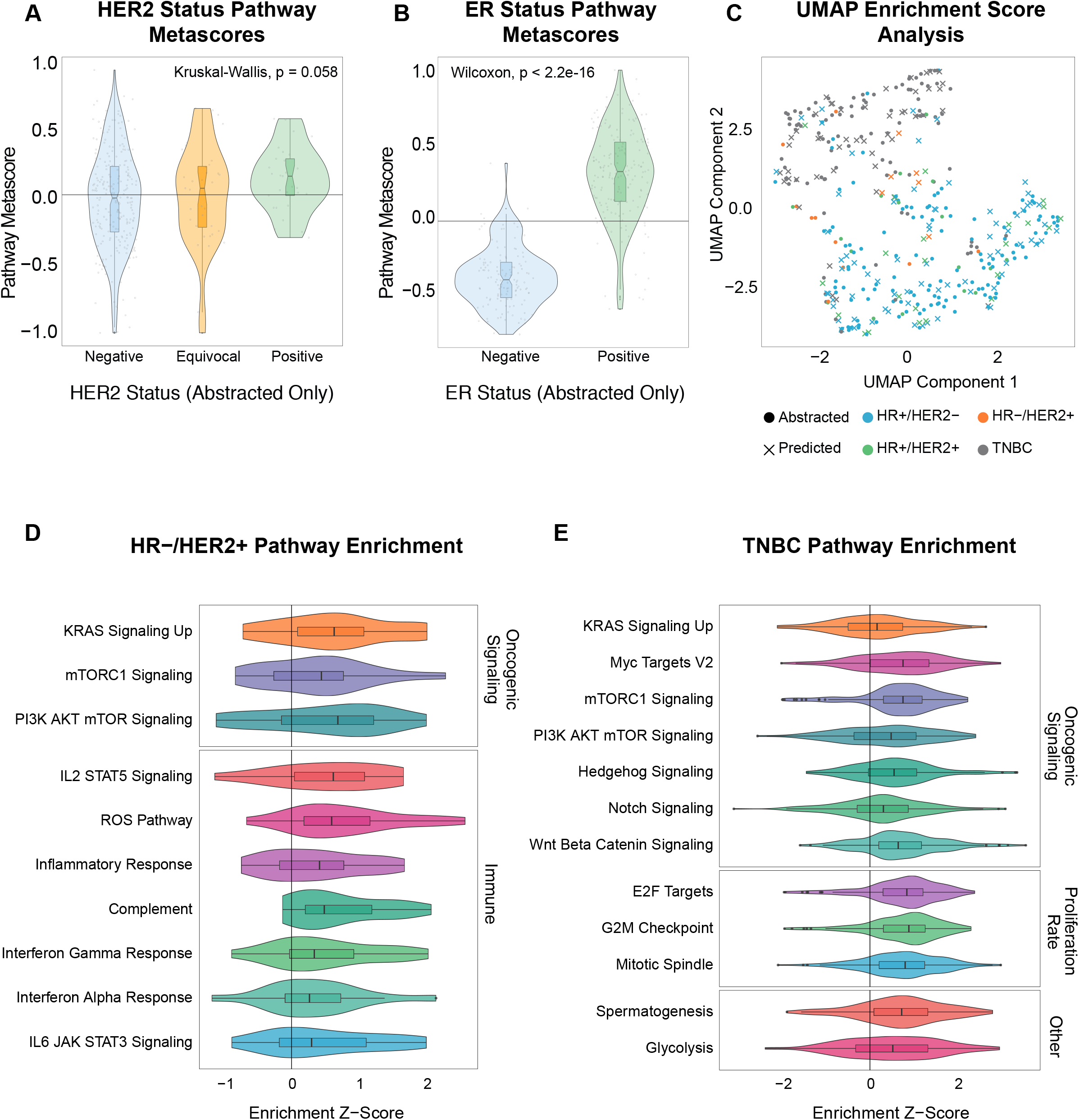
RNA-seq breast cancer pathway analyses of the Tempus molecular sequenced cohort. (A) HER2 and (B) ER pathway metascores for patients with abstracted HER2 IHC or FISH test results. (C) UMAP of 50 Hallmark enrichment scores. Patients with molecular subtypes based on at least one abstracted receptor status are depicted by circles, while patients with molecular subtypes determined exclusively from RNA-predicted statuses are depicted by X symbols. Distribution of enrichment Z-scores for (D) HR-/HER2+ and (E) TNBC relevant pathways.

Next, RNA-seq data were analyzed in relation to the highly curated Hallmark pathway gene sets to determine the differential activation of biological pathways between breast cancer subtypes.^45^ Most Hallmark pathways (32 of 50) exhibited significantly different enrichment scores between molecular subtypes (**Supplemental Fig. 4A**). A UMAP using only scores from these 50 pathways recapitulated the TNBC clustering observed in the full-transcriptome UMAP (**Fig. 6C**). As expected, HR+ samples, but not HR- or TNBC samples, were highly enriched for two pathways related to estrogen signaling (**Supplemental Fig. 4B**). Among HR-/HER2+ cancers, we observed enrichment for pathways known to be downstream of HER2, RAS, and mTOR (**Fig. 6D**).^63^ HER2-driven tumors also showed enrichment for all immune-related Hallmark pathways, a finding consistent with the literature.^64^ Many oncogenic signaling pathways were enriched in TNBC (**Fig. 6E**), including Wnt, mTOR, PI3K, Hedgehog, and Notch, consistent with TNBC tumors’ reliance on ER-, PR-, and HER2-independent pathways.^65^ TNBC samples were also enriched for pathways related to mitotic index, as expected due to their relatively high growth rate,^66^ glycolysis, which is consistent with their elevated Warburg effect and potentially targetable,^67^ and cancer/testis antigens.^68^

## Discussion

The expanding utility of RWE is evident with the growing number of related studies and regulatory considerations.^2,3,7,69^ Compared with randomized controlled trials, however, RWD analyses are complicated by a lack of standardization between records and the introduction of extraneous factors, such as natural language processing errors and uncontrolled confounding variables.^2,3,7,22-24^ We aimed to address these concerns by 1) increasing the statistical power of analyses with a relatively large cohort size, 2) incorporating a variety of data sources beyond electronic health records to benefit downstream analyses,^1,3,22,27^ and 3) demonstrating consistency between characteristics of the real-world cohort and results from previous clinical studies.

Using only a portion of breast cancer patient records from the extensive Tempus clinicogenomic database, our retrospective analysis provides further evidence for the feasibility and value of generating clinically relevant RWE. We first demonstrated that longitudinal RWD can capture key information regarding patient clinical history, treatment journey, and outcomes. Our RWD analyses generated valid RWE that replicated previously published clinical results and was generally consistent with established databases, indicating feasibility. Although the majority of cohort characteristics were aligned with previous clinical studies, the analyses also highlighted the complexities in breast cancer RWD. For instance, the proportion of pre- and post-menopausal patients was similar to previous clinical trial data,^49^ but menopausal status was only confidently abstracted in approximately 51% of the cohort. Upon further review, many RWD breast cancer studies have either applied simplified definitions of menopause, such as an age cutoff,^19^ reported missing statuses in electronic records,^8,70^ or did not include menopausal status at all. Simplifying rules for abstraction may fill these gaps in RWD, such as in defining real-world progression-free survival, but can also affect the validity of conclusions.^71,72^

To strengthen the validity of RWE presented here, rules were established and applied to perform relevant analyses and derive statistics from the cohort. For example, rules described in the methods facilitated the definition of molecular subtypes from multiple abstracted test results. Our HER2 test result analyses confirm the existing conflict in standard testing interpretations, an issue evident by recent American Society of Clinical Oncology (ASCO) guidelines, previous clinical studies, and meta-analyses.^54,73-76^ Specifically, our findings of IHC intra-test discordance illustrate the subjectivity of IHC testing, prompting standard testing improvements and biomarker discovery.

Upon observation of discrepancies in abstracted HER2 testing results, a separate cohort with complete biopsy data was selected to test the efficacy of a whole-transcriptome model in predicting molecular subtypes. By combining clinical and molecular data, we demonstrate transcriptome profiling is complementary to RWD and can illuminate fundamental biological differences between patients. RNA-seq may supplement standard testing interpretations by providing clinically relevant insights when biopsy test data is inconclusive, exemplified here in the resolution of molecular subtypes for patients with equivocal statuses. Addressing these cases is critical in the evolving treatment landscape, as molecular subtypes are key criteria for breast cancer treatment decisions.

Furthermore, our signaling pathway investigation uncovered potential pathway-related therapeutic targets, such as oncogenic signaling via the mTOR pathway, for subtypes like TNBC with limited pharmacotherapies available. RNA pathway analyses can also elucidate treatment-related tumor characteristics not captured by standard diagnostic and prognostic tests, such as additional biomarkers or amplifications that may be targetable in HER2+ breast cancer patients.^77-79^ Expression-based immune signatures can also predict response to neoadjuvant treatment with several experimental agents/combinations added to standard chemotherapy, including the addition of pembrolizumab in early-stage TNBC.^80^ Biomarker selection of immunotherapy in early-stage TNBC will become imperative to therapeutic strategies given its substantial toxicity.

## Conclusion

The Tempus data pipeline integrates longitudinal RWD and comprehensive molecular sequencing data into a structured clinicogenomic database capable of generating valid clinical evidence in real-time. While RWD are inherently complex, cancer cohort selection and data insights are feasible using structured data sources and strictly defined analysis criteria. Finally, integrating RNA-seq data with RWD can improve clinically actionable evidence related to clinical markers, potential therapeutic targets, and optimal therapy selection in breast cancer.

## Clinical Practice Points

- The feasibility of real-world data (RWD) analysis has increased alongside technological advances and regulatory support to continuously capture and integrate healthcare data sources. Several studies demonstrate the ability for real-world evidence (RWE) to guide clinical development strategies, expand product labels, and address knowledge gaps by examining clinical aspects not captured in clinical trials.
- Despite recent advances and growing regulatory support, RWD from heterogenous structured and unstructured sources is often challenged by various technical barriers. Lack of standardization between electronic records, underpowered natural language processing tools, and uncontrolled extraneous variables threaten the validity of well-sourced RWE.
- Our RWD analyses followed strict qualitative criteria to produce RWE of demographics, clinical characteristics, molecular subtype, treatment history, and survival outcomes from a large, heterogeneous database. Importantly, the results were mostly consistent with data from previous clinical studies, suggesting feasibility of generating valid RWE. We also demonstrate the value of integrating omics data with RWD through the use of whole-transcriptome analyses in relevant breast cancer signaling pathways and a predictive model for receptor statuses.
- These data provide rational for use of the Tempus clinicogenomic database to generate RWE and conduct real-time, hypothesis-driven analyses of large RWD cohorts in the future. Clinicians may utilize these large-scale databases to circumvent the restrictive exclusion criteria of controlled studies, clarify real-world patient needs, and aid the development of clinical trials. Furthermore, our results suggest molecular data may bolster deficiencies in standard breast cancer diagnostic tests.

## Data Availability

Relevant data are available within the text, figures, tables, and supplementary information.

## Acknowledgments

We are thankful to ASCO CancerLinQ for their partnership and the clinical data, operations, data science, engineering, pathology, and lab teams at Tempus Labs. We sincerely thank the entire clinical data abstraction team, Jeff Ottens, and April Manhertz. We thank Kelly McKinnon for proofreading and figure assembly and design. We thank Kevin White and Kimberly Blackwell for scientific review and discussion of the manuscript. We thank Hailey Lefkofsky for initial discussions and Eric Lefkofsky for his support and discussions.

Supplemental Figure and Table Legends

**Supplemental Figure 1**.Patients grouped by year of initial diagnosis. The distribution of patients by year of initial diagnosis across the clinical abstraction cohort.

**Supplemental Figure 2**.Molecular characteristics of the Tempus molecular sequenced cohort. (A) The distribution of patients with variants in the most frequently reported genes across the cohort. The number of patients harboring mutations in each gene are shown above the bars. (B) The number of variants classified as alterations, amplifications, or deletions within each of the most frequently reported genes in the cohort. (C) The distribution of patients with pathogenic germline alterations in NCCN-designated familial high-risk genes and (D) TMB across the cohort.

**Supplemental Figure 3**.Breast cancer pathway analyses from RNA-seq data of the Tempus molecular sequenced cohort according to MSigDB and Hallmark pathways. (A) Pearson correlation between *ERBB2* expression and enrichment scores (GSVA) for each HER2-related pathway in MSigDB among the cohort. (B) Correlation between *ESR1* expression and enrichment scores for each ER-related pathway in MSigDB among the cohort.

**Supplemental Figure 4**.(A) For each Hallmark pathway, the significance of differential enrichment between molecular subtypes was determined by a Kruskal-Wallis test of the enrichment scores. The vertical line indicates *P*=0.001 and any value to the right of the line was considered significant. (B) Distributions of z-scores among HR+/HER2- (blue), HR+/HER2+ (green), HR-/HER2+ (orange), and TNBC (grey) patients for the two estrogen response Hallmark pathways with the most significant differential enrichments between molecular subtypes.

**Supplemental Figure 5**.Distribution of enrichment z-scores for each HER2-related pathway in MSigDB among patients in the Tempus molecular sequenced cohort. Patients with negative (blue), equivocal (orange), or positive (green) abstracted or predicted HER2 test results are shown. The *P*-values listed for each pathway represent the results of a Kruskal-Wallis test for the difference between enrichment scores from HER2-, HER2-equivocal, and HER2+ patients.

**Supplemental Figure 6**.Distribution of enrichment z-scores for each ER-related pathway in MSigDB among patients in the Tempus molecular sequenced cohort. Patients with negative (blue) or positive (green) abstracted or predicted ER test results are shown. The *P*-values listed for each pathway represent the results of a Wilcox rank sum test for the difference between enrichment z-scores from ER+ and ER-patients.

**Supplemental Table 1**.Inter-test comparison of HER2 status from IHC and FISH results among patients in the clinical abstraction cohort with both tests conducted at initial diagnosis.

**Supplemental Table 2**.Single-gene logistic model performance results for RNA-based predictions of ER, PR, and HER2 status in the Tempus molecular sequenced cohort.

